# ‘It’s about dying, not just a broken leg’ - Qualitative findings on paramedics’ perception of end-of-life situations in rescue scenarios in Germany

**DOI:** 10.1101/2023.01.11.23284420

**Authors:** Nicola Rieder, Christian Banse, Franziska Schade, Friedemann Nauck

**Author notes:** Corresponding author: Nicola Rieder M.Sc., Department of Palliative Medicine, University Medical Center Goettingen, Von-Siebold-Str. 3, 37075 Goettingen.

## Abstract

In Germany, paramedics and emergency physicians arrive separately on scene (‘rendez-vous-system’), which aims to save resources when no physician is needed. Usually, paramedics arrive firstly on scene, and are obliged by law to perform all possible action to maintain a patient’s life. Especially in end-of-life (EoL) situations, this can cause conflicts, as those patients may require a decent consideration between ‘saving life’ and ‘allowing to die’. Whereas the emergency physicians’ perspective on this issue is relatively well examined, until today, in Germany, little is known about the (non-physician) paramedics’ perception.

**Aim:** To explore paramedics’ perception of rescue scenarios involving patients with advanced, incurable, severe diseases facing their EoL and to scientifically classify this cross-sectional field through experts from different research fields.

**Method:** Seven semi-structured narrative interviews with paramedics and one focus group with representatives from the Center for Medical Law in Goettingen were conducted and analysed using qualitative content analysis.

**Results:** Six key categories emerged from the data. Rescue scenarios in EoL situations are perceived as highly complex situations that are narrowed down through deviating goals of care (1), limited options for action (2), an emotional overload of all involved actors (3), consequences of a neglected (care) planning in advance (4) and various background structures, such as current societal and demographic changes (5) and systemic challenges (6). Complexity also arises from the multiple influences of the different categories/ their content on one another.

**Discussion:** Being confronted with patients that do not wish for further life-maintaining treatment marks a significant shift concerning the range of rescue scenarios, resulting from (i.a.) demographic developments and structural dynamics in health care. Therefore, in addition to specific actions restoring paramedics’ ability to act in rescue scenarios in EoL situations, a general discussion of the emergency services’ area of responsibility as well as the (emergency) medical treatment and care of patients with advanced, incurable, severe diseases is required.

## Background

Emergency cares’ initial scope is to provide urgent pre-hospital treatment and stabilisation for serious illnesses and injuries, and transport patients to definitive care. Over time, the number of patients in prehospital emergency care has been noticeably increasing, resulting not only from an increase in life-threatening events in total, but also a general increase in the use of emergency care (1,2). Therefore, emergency care has been in the centre of health policy attentions and discussions around efficiency and effectiveness for several years (3–7). Nowadays, pre-hospital workflows are characterised by algorithms to structurally and effectively process different rescue scenarios. These algorithms standardise the entire operational process through explicit guidelines and instructions, from arriving on scene until transferring the patient to hospital care (8). Previously, inconsistent standards of care repeatedly led to scientifically proven deficits in preclinical patient care (9,10). In Germany, prehospital emergency care implies a “rendez-vous-system” where paramedics and emergency physicians approach separately on scene (11), aiming to save resources in rescue scenarios where no emergency physician is needed. In addition, two professional levels of paramedics regulated by the federal law exist: “Rettungsassistent*in” (two year education, effective from 1989 until 2013) and “Notfallsanitäter*in” (three-year education, effective since in 2014), both are able to provide first level pre-hospital emergency care. In most instances, paramedics arrive first on scene. One major disadvantage of the rendez-vous approach may be a delayed arrival or a lack of availability of the emergency physician personnel when its urgently needed, particularly in situations where “maintaining life” does not necessarily equals the provision of goal-concordant care.

This might be the case, when patients’ best possible physical recovery is generally limited: Expectations, wishes and needs of patients with advanced, incurable, severe diseases increasingly gear towards maintaining the best possible quality of life rather than maintaining life itself. In those situations, the indication of invasive or even intensive care treatments seem increasingly questionable (12,13). This rather “palliative” understanding competes with the principle of emergency care, to undertake all effective measures and treatments to restore and maintain a patient’s vital functions.

Research evidence shows that those situations can be extremely distressful for both, physician and (non-physician) paramedic personnel (14,15). Whereas the emergency physicians’ perspective is relatively well examined (16,17), until today little is known about paramedics’ perception of those situations. Since paramedics in Germany are principally not permitted to modify therapy goals and/or decide independently on limitation of therapies (as this right is reserved to physicians), they are legally obliged to perform all possible and indicated action to maintain a patient’s life (18). We suspect that this may lead to complex quandaries, as from paramedics’ experience, comfort care might be more appropriate than aggressive, interventional procedures.

Furthermore, a scientific evaluation of this cross-sectional field (e.g. a combination of medical, legal and ethical perspectives) is missing in Germany.

## Aim

In this study, we aimed to explore i) how (non-physician) paramedics perceive rescue scenarios involving patients with advanced, incurable, severe diseases, facing their end of life in Germany, ii) which support or change is desired and iii) how experts from different research subjects (here: representatives from the Center for Medical Law in Goettingen) evaluate the situation.

### Material and Methods

We used qualitative research methods to assess and interpret the matter. To ensure a high scientific standard and transparency, the reporting of this study will follow the “Consolidated criteria for reporting qualitative research” (COREQ) Checklist (19).

### Recruitment

Data were collected between March 2020 and July 2020.

We used semi-structured (guided) interviews to explore paramedics’ experiences and the meanings they attribute to them. Additionally, we conducted an online focus group at the Center for Medical Law in Goettingen, whose main task is the scientific research regarding current questions and problems that have arisen in those areas, where Medicine, Ethics in Medicine and Law collide.

Paramedics (n=7) were recruited using existing contacts to study participants of a previous research project (15) as well as using NRs’ personal contacts to intermediaries in the target group such as Heads of Division in rescue services and/or Chief Fire Officers of Fire and Rescue Services resulting from her own work experience as a paramedic. Care was taken to select the participants as heterogeneously as possible regarding their areas of application.

Participants were contacted via Email and phone and were informed about the study, its aims and conduct. Participants should have experienced at one conflictual EoL situation while working in emergency service and must be willing to talk about it.

Furthermore, an online focus group with representatives from the Center for Medical Law in Goettingen was conducted (20). Five experts from different subjects were asked to discuss the preliminary results in order to assist in structuring, categorising and interpreting them. NR and FN actively contacted representatives from the following areas: Criminal Law, Medical Law, Palliative Medicine, Systematic Theology and Ethics in Medicine. The participants were contacted via Email and phone and were informed about the study, its’ aims and conduct.

If participants were interested to participate in the interviews, the study information including data management procedures was sent via mail. Participants received an informed consent form, and sent it back via mail.

### Data collection

NR prepared an interview guide and discussed the content in the group of the co-authors (FS, CB, FN). The final guide contained the following questions/ narrative prompts:

Please tell me about at least one self-experienced rescue scenario with patients with advanced, incurable, severe diseases facing their end of life.

Which challenges did you meet? Where were the differences to ‘regular’ rescue scenarios?

How did you manage those challenges? Please tell me about the strategies and solutions that worked best for you.

What strategies and solutions would you choose again to deal with such situations. Where do you need more support?

Participants (n=7) were encouraged to talk about issues pertinent to the research questions in a one-to-one interview by NR. The interviews were recorded on a Dictaphone. Due to Covid-19 contact restrictions, NR conducted the interviews by phone. NR is a public health researcher, with a focus on the interface between palliative care and emergency medicine. She has worked as a paramedic. She has experience in conducting interviews and (online) focus groups and works at the Department of Palliative Medicine at the University Medical Center Goettingen in a research project that focuses on Advance Care Planning (ACP).

For the online focus group, NR adapted the interview guide to the preliminary results. Beforehand, NR presented them to the participants, to give them the opportunity to discuss the topics.

Due to Covid-19 contact restrictions NR conducted the focus group online using Big Blue Button, a software that is locally hosted by the University Medical Center Gottingen’s computing center, which ensured conformation with data security regulations. NR was supported by FS. The focus group was recorded with an inbuilt recorder (video and audio) and, additionally, on a dictaphone (audio only).

During the focus group, we used knowledge mapping to create a focus group illustration map (21). With this method, a researcher records results as a ‘mind map’, which can be discussed with the participants to validate results, and ensure that participants’ statements were understood. We discussed the knowledge map with the focus group at the end of the discussion. Two researchers took part in the focus group. NR moderated the focus groups and FS made notes and prepared the focus group illustration maps during the focus group. FS is a sociologist and public health researcher, with experience in conducting focus groups with health care providers in palliative care. She also works as a researcher at the Department of Palliative Medicine at the University Medical Center Goettingen. At the beginning of the focus group, researchers and participants introduced themselves.

Both, interviews and the focus groups statements were transcribed verbatim.

### Data analysis

Both interviews and the online focus group were analysed using qualitative content analysis (22) to systemically organise the data into a structured format. To prepare the focus group, interviews were analysed first. NR inductively coded each interview, and grouped codes to form key categories, using MAXQDA Analytics Pro 2018. Codes and categories were discussed with FS and CB. Then, these categories were used to deductively code the focus group. Codes that did not fit into the existing categories were brought together to form new categories. During the coding, various associations and dependencies between categories emerged, which are outlined in Figure 1.

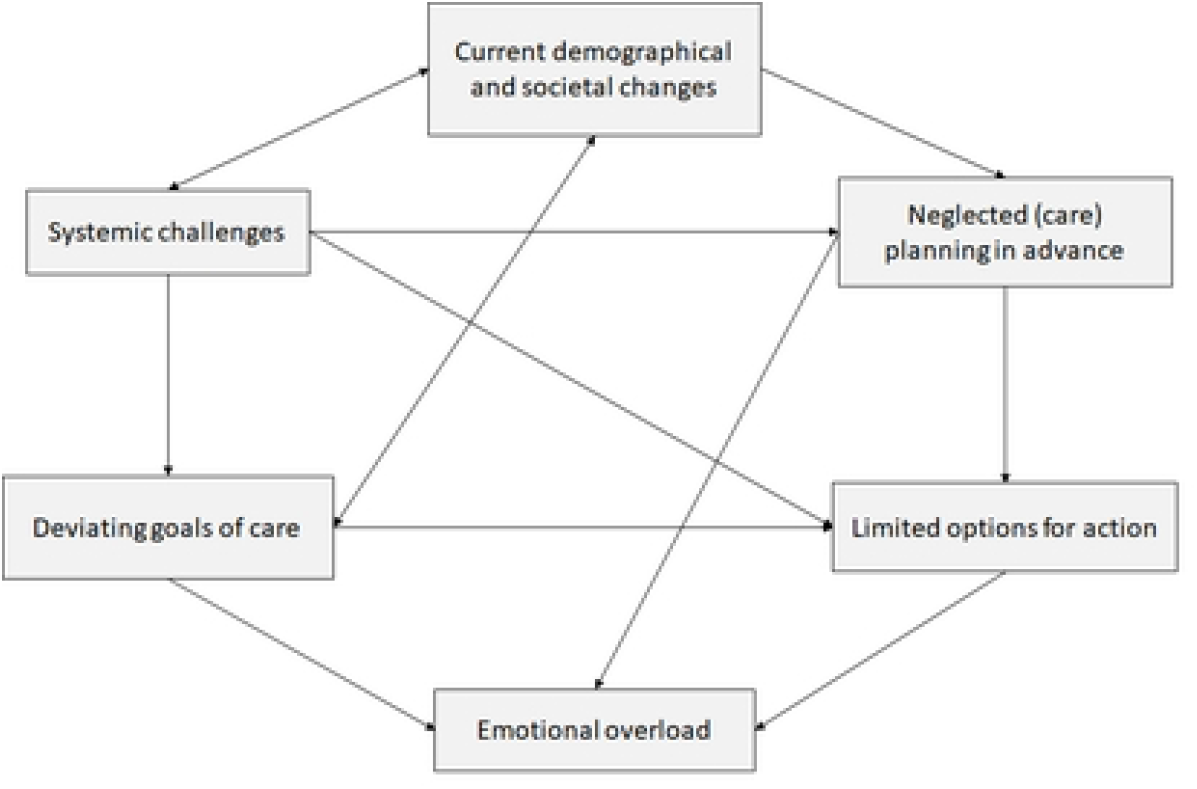

### Ethics

The study received approval from the Research Ethics Committee of the University Medical Center Goettingen (21/10/19).

## Results

### Sample

Between June 10, 2020, and July 08, 2020, seven interviews were conducted. The interviews lasted between 59 minutes and 1 hour and 21 minutes. Participants (age range: 27 - 42 years) were working full-or part-time as a paramedic (“Rettungsassistent*in” (n=4) or “Notfallsanitäter*in” (n=3), *Table 1*), with a total work experience between five and 22 years. Two interviewed paramedics were female and five were male. Three had an additional qualification as practical instructors. Three participants were qualified as rescue service organisational commanders. Care was taken to select the participants as heterogeneously as possible regarding their areas of application, due to differing organisational structures, e.g. medical treatment standards that are specified by the competent medical management, or the thematic design of further education. Participants were recruited from five different rescue stations in four different federal states of Germany.

**Table 1:**
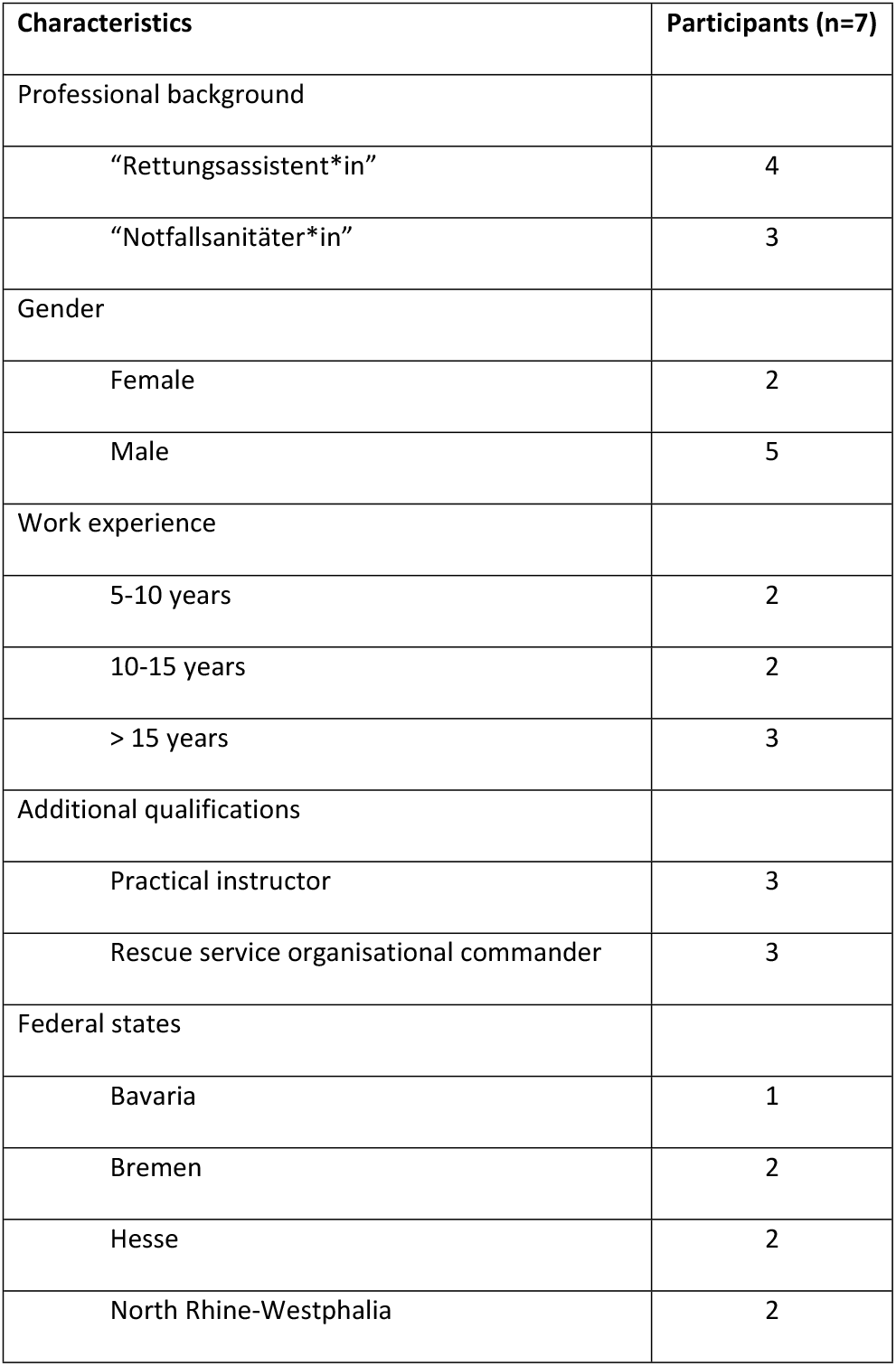
Participant characteristics (interviews).

| Characteristics | Participants (n=7) |
| --- | --- |
| Professional background |  |
| “Rettungsassistent*in” | 4 |
| “Notfallsanitäter*in” | 3 |
| Gender |  |
| Female | 2 |
| Male | 5 |
| Work experience |  |
| 5-10 years | 2 |
| 10-15 years | 2 |
| > 15 years | 3 |
| Additional qualifications |  |
| Practical instructor | 3 |
| Rescue service organisational commander | 3 |
| Federal states |  |
| Bavaria | 1 |
| Bremen | 2 |
| Hesse | 2 |
| North Rhine-Westphalia | 2 |

The focus group lasted 1 hour 54 minutes, including the content-related discussion of the focus group illustration map. Participants were representatives from the following areas of research: Criminal Law, Medical Law, Palliative Medicine, Systematic Theology and Ethics in Medicine. All participants (n=5) of the focus group were male.

### Key categories

A total of six key categories emerged from the data. *Fig 1* gives an overview of the categories and shows their relationships to each other. To support our interpretation of the data, we provide quotations to illustrate the categories in *Table 2*.

### Deviating goals of care

Emergencies are commonly characterised by a distinct existentiality and the limited availability of resources such as equipment, time and knowledge about patients’ situation or medical history. Although those aspects generally shape all emergencies, diverging goals of care and the non-transferable individuality that is contrasting the paramedics’ usual guideline-based way of work are strongly contradicting in rescue scenarios in EoL situations. Additionally, paramedics report the feeling that enabling dying somehow differs from their common self-perception as rescue personnel.

### Limited options for action

Paramedics describe great uncertainties regarding legal foundations and lawful acting when neglecting life-sustaining treatment and not only considering death as a possible (negative) outcome but eventually enabling dying. This contributes to a willingness of the paramedics to pass and delegate those decisions to superior structures (emergency physicians, hospital etc.), leading to a diffusion of responsibilities. Furthermore, a lack of education and frameworks when dealing with patients with advanced, incurable, severe diseases at their EoL, and the wish for intra- and interprofessional content-related training, further education on advanced, incurable, severe diseases, retrospective case conferences/ professionally guided medico-ethical reflection and any kind of (legal) framework to manoeuver the situation is described. Nonetheless, the introduction of a “palliative algorithm” is viewed critically, as paramedics doubt that the individuality of one’s own death and dying can be translated in one universal guideline. Another aspect emphasises the general lack of rescue services’ access to and networks with geriatric and/or palliative departments that limits options for the initiation of alternative care.

### Emotional overload

The existentiality of the situation and the lack of preparation and awareness result in an emotional overload of all involved actors (e.g. paramedics, relatives, caregivers). All paramedics report a distinct feeling of being overwhelmed, when confronted with situations, where their “normal” (life-maintaining) approach is neither suitable nor wished for and seems out of place, resulting in feeling helpless, not being able to take charge of the situation and rather being at its mercy. In addition, relatives or bystanders tend to pass their emotional overload and existential decisions unfiltered on to the paramedics, expecting them to tell them what to do and decide. Paramedics perceive this as strongly demanding and overstraining, strengthening their fear of doing something wrong. According to the focus groups’ discussion, the latter might be a product of the discordance between rescue services’ mandate to perform all possible action to rescue and maintain life and the deviating goals of care in EoL situations, where further life-maintaining treatment is not necessarily prioritised.

### Neglected (care) planning in advance

From paramedics’ perspective, rescue scenarios in EoL situations reveal both, importance and the absence of preparation in terms of accessible, valid, meaningful, ascertainable and updated advance directives, and the communication of their content to the authorised representatives or proxies. This leads to confusion and insecurities amongst paramedics when confronted with those documents, resulting in an attitude that is described as “rather carry them to hospital and feel safe”. Paramedics also agree on a lack of awareness regarding both, the actual health status and the debate about the approaching death as well as its implications by patients, family members, relatives, caregivers etc. as a possible trigger for conflictual situations. Hence, the dissolving of societal taboos on death and dying and instead an open debate on it seems necessary to them. Family carers’ knowledge about the implications of the preference to die at home is limited. Also, the availability of willing and able family or professional carers and the willingness and ability of general practitioners and/or specialised palliative home care teams to visit and support patients at home is limited. In addition, the general practitioners’ (GPs) role also represent an important aspect to the paramedics. From their perspective, GPs can/should play a key role in the supply of early, understandable information about the illness situation and health status, and can/should open the doors to appropriate and necessary care, but are perceived (by paramedics) as rather reluctant when it comes to breaking bad news and advance care planning.

### Current demographic and societal changes

Both, interviewees’ and the focus group participants’, intensively discussed the implications of current demographic and societal challenges, such as the lack of societal health education and the increase in chronically ill, multimorbid and/or terminally ill patients in contrast to the health care systems’ lack of structural adaptation. There is consensus that health education and an effective publicly communication of other appropriate and accessible contact structures and persons in those situations are needed.

### Systemic challenges

The questionable concordance between emergency services’ mandate and deviating goals of care in EoL situations contribute to the fact that high demands and expectations meet a somehow “restrained” health system. Inconsistencies and unanswered questions in what paramedics can or cannot do (in Germany, the Federal States are in charge of the emergency-service-specific legislation), how to cope with consequential differences in competencies, and how this “catalogue of service” is communicated to the general public, causes confusion on various levels. Additionally, distinct hierarchies evolving over the past decades are making the power disparity between physician and non-physician personnel very clear. According to the focus groups’ discussion, systemic complexity, e.g. knowing when to contact or reach out to which health care provider (general practitioners, on-call medical services, emergency rescue services, specialised palliative home care) is another factor strengthening complexity. According to the focus groups’ discussion a political reflection and clarifications of the emergency services’ function, objectives and responsibilities is strongly advised.

## Discussion

The confrontation with patients at their EoL, who do not wish for further life-maintaining treatment marks a perceived shift concerning the range of rescue scenarios. Rescue scenarios in EoL situations are perceived as highly complex situations that are narrowed down through deviating goals of care, limited options for action, an emotional overload of all involved actors, consequences of a neglected (care) planning in advance, and various background structures, such as current societal and demographic changes and systemic challenges. Complexity also arises from the multiple influences of the different categories on one another.

The increase in terminally ill patients associated with demographic change is amplified by a lack of early integration into appropriate care structures. This observation is consistent with a recent study (23), examining hospitalisation rates and causes of nursing home residents. Above all, especially non-oncological, terminally ill patients (geriatric, dementia, multimorbid and/or chronically ill patients) often fall through supply networks and are consequently caught repeatedly by emergency care structures. Access and connection to palliative care (esp. specialised palliative home care) is still limited, for both patients and rescue service, due to bureaucratic obstacles. Clarification who can be a reliable, main contact person for those patients, what is required in terms of patient information and education as well as pro re nata (PRN) medication and how this information can be communicated to all involved actors, is needed. Hence, bolstering grass-roots health structures at a political and (healthcare) systemic level, and an effective, more centralised coordination of requests for help that can directly contact e.g. palliative care providers seems to be indispensable. Those aspects are part of legislative endeavours to reform the emergency care (9).

An open dialogue, educational offerings and the promotion of regional networks are key issues for a profound literacy in aspects of death and dying. In addition to the assumption that this is the responsibility by GPs, which was discussed by the interviewees, low-threshold information and education for citizens (e.g. “Letzte Hilfe Kurse”) enable them to talk openly about death, dying and care at the end of life, and contribute to the regional networking of formal and informal support services (24).

Additionally, concepts for a better preparation of the paramedics, such as specifically (geriatric) trained paramedics or specific geriatric emergency departments that meet the needs of aging, multimorbid patients, are needed (25,26). Regarding the implementation of specific algorithms, Makowski et al. published a guideline in 2013 for individual decision-making for palliative patients addressing the physician emergency personnel, which is based on a categorisation of palliative emergency situations. It aims to present possible options for action (27). Further research should discuss a possible transferability to the paramedics’ situation.

Paramedics reporting their helplessness and feelings of being out of place when being confronted with patients that do not wish for further life maintaining treatment mark a vital concern for further research and possible adjustments of emergency care’ scope and paramedics’ training and education. The assumption that there are worse things than death and dying may reflect on paramedics’ fear to enhance harm by prolonging suffering instead of help and rescue when being onsite. Therefore, a reflection on (moral) expectations paramedics (should) have seems important. According to the literature, those aspects could be met by specific education and training, including guided (self-)reflection on experienced situations or case studies, terms and principles of medical ethics and law, paramedics’ professional self-image, EoL care, and methods of moderated problem-solving (28).

Paramedics report an urgent need and wish for standardised, comprehensible and valid advance directives, which offer the opportunity for an acceptable and (medical) legal framework for all involved parties. “Advance Care Planning” (ACP) serves as an example for this: it refers to a comprehensive consulting and implementation concept in which individual informational and conversational interventions to empower individuals are combined with the implementation of regionally uniform crisis and emergency forms that is carried out on an institutional and systemic level of all relevant care structures -including emergency structures (29).

### Limitations

Due to the Covid-19 contact restrictions and the limited period of the study, we conducted the interviews via phone and the focus group online as a video conference. Therefore, non-verbal communication was limited (30). While this potentially restricted the interviewees’ and the group’s interaction, we judged this as acceptable as our research interest focused on their experiences. The non-“face-to-face” -situation allowed us to rapidly recruit participants that were geographically dispersed and had – especially concerning paramedics’ high workload during the beginnings of the Covid-19 pandemic - limited time resources.

The use of qualitative content analysis was and is suitable regarding our research question and our research focus on explicit knowledge. Nonetheless, limits of this method moving at the surface of a research problem became apparent: questions about causal conditions e.g. motives for certain behaviours, or conditions intervening the designated strategies remain unanswered. As described in the results, categories and sub-categories are interconnected and mutually influence each other. According to the main category, the issue seems too complex to be represented exclusively with the possibilities of qualitative content analysis. Our results illustrate, that the identified issues might be influenced by e.g. aspects of one’s own (professional) attitudes and motivation, the individual and societal perception of death and dying, and the societal understanding of the emergency carers’ service mission. Furthermore, bringing together different perspectives (patients, relatives, other caregivers) might positively contribute to a comprehensive understanding. In addition, fundamental debates about the organisation, tasks and responsibilities of emergency services in Germany, and about the medical, social and ethical perception of aging and dying people are touched. The effect of new diagnostic options and modern intensive care medicine and its consequences, as well as the implications of empowering patients’ autonomy and a (perceived) lack of health and death literacy, were not considered in detail. Further research that goes methodically in-depth is necessary, especially because the data cannot be generalised.

## Conclusion

The aim of our research was a better understanding of the various aspects of conflictual situations experienced by paramedics in EoL rescue scenarios, to collect and derive knowledge about possible approaches to solutions and their location on different levels, in different areas and with different stakeholders. We learned that paramedics in Germany perceive rescue scenarios involving patients with advanced incurable, severe diseases facing their EoL as highly complex and that they take over an important role they do not feel well prepared for.

## Data Availability

Data cannot be shared publicly because of limited anonymity. Data are available from the Department for Palliative Medicine at the University Medical Center Goettingen (contact via) for researchers who meet the criteria for access to confidential data.

## Acknowledgement

We would like to thank all study participants for contributing their time and experiences.

## Author Contributions

Conceptualisation: Nicola Rieder, Christian Banse, Friedemann Nauck

Data curation: Nicola Rieder, Franziska Schade, Christian Banse.

Formal analysis: Nicola Rieder, Franziska Schade, Christian Banse.

Funding acquisition: Friedemann Nauck

Investigation: Nicola Rieder.

Methodology: Nicola Rieder, Franziska Schade, Christian Banse.

Project administration: Nicola Rieder.

Supervision: Friedemann Nauck.

Validation: Friedemann Nauck.

Writing – original draft: Nicola Rieder.

Writing – review & editing: Nicola Rieder, Franziska Schade, Christian Banse, Friedemann Nauck.

## References

1. Seeger C. Interdisziplinäres Arbeiten im Team — Grundlage für die Vernetzung von Palliative Care. In: Palliative Care [Internet]. Berlin, Heidelberg: Springer Berlin Heidelberg; 2012 [zitiert 7. März 2012]. S. 171–84. Verfügbar unter: http://www.springerlink.com/index/10.1007/978-3-540-72325-7_11.

2. Hobbs FDR, Bankhead C, Mukhtar T, Stevens S, Perera-Salazar R, Holt T, u. a. Clinical workload in UK primary care: a retrospective analysis of 100 million consultations in England, 2007–14. The Lancet. 4. Juni 2016;387(10035):2323–30.

3. Gries A, Bernhard M, Helm M, Brokmann J, Gräsner JT. Zukunft der Notfallmedizin in Deutschland 2.0. Anaesthesist. Mai 2017;66(5):307–17.

4. Gretenkort P, Beneker J, Dörges V, Fischer L, Kann D, Sefrin P. Strukturänderungen in der präklinischen Notfallmedizin - Standortbestimmung 2016. Notarzt. 20. Dezember 2016;32(06):264–70.

5. Fachexperten der Eckpunktepapier-Konsensus-Gruppe, Fischer M, Kehrberger E, Marung H, Moecke H, Prückner S, u. a. Eckpunktepapier 2016 zur notfallmedizinischen Versorgung der Bevölkerung in der Prähospitalphase und in der Klinik. Notfall Rettungsmed. August 2016;19(5):387–95.

6. Sefrin P. Neuordnung der Notfallversorgung im ambulanten/präklinischen Bereich. NOTARZT. Juni 2018;34(3):132–9.

7. Durand AC, Gentile S, Devictor B, Palazzolo S, Vignally P, Gerbeaux P, u. a. ED patients: how nonurgent are they? Systematic review of the emergency medicine literature. The American Journal of Emergency Medicine. 1. März 2011;29(3):333–45.

8. Peters O, Runggaldier K, Schlechtriemen T. Algorithmen im Rettungsdienst: Ein System zur Effizienzsteigerung im Rettungsdienst. Notfall Rettungsmed. Mai 2007;10(3):229–36.

9. Messelken M, Dirks B. Zentrale Auswertung von Notarzteinsätzen im Rahmen externer Qualitätssicherung. Notfall & Rettungsmedizin. 1. Oktober 2001;4(6):408–15.

10. Schlechtriemen T, Lackner ChrK, Moecke Hp, Stratmann D, Altemeyer KH. Sicherung der flächendeckenden Notfallversorgung: notwendige Strukturverbesserungen. Notfall & Rettungsmedizin. 1. Oktober 2003;6(6):419–28.

11. Sempere-Selva T, Peiró S, Sendra-Pina P, Martínez-Espín C, López-Aguilera I. Inappropriate use of an accident and emergency department: Magnitude, associated factors, and reasons—An approach with explicit criteria. Annals of Emergency Medicine. Juni 2001;37(6):568–79.

12. Alt-Epping B, Geyer A, Nauck F. [Palliative care concepts for patients with non-oncological diseases]. Dtsch Med Wochenschr. August 2008;133(34–35):1745–9.

13. Nauck F, Alt-Epping B. Crises in palliative care—a comprehensive approach. The Lancet Oncologist. 2008;9:1086–91.

14. Smith AK, Fisher J, Schonberg MA, Pallin DJ, Block SD, Forrow L, u. a. Am I Doing the Right Thing? Provider Perspectives on Improving Palliative Care in the Emergency Department. Annals of Emergency Medicine. Juli 2009;54(1):86-93.e1.

15. Rieder N, Mühe K, Nauck F, Alt-Epping B. “Leben retten bis der Arzt kommt?”: Konflikte und wahrgenommene Belastung des nichtärztlichen Rettungsdienstpersonals im Umgang mit Patienten mit fortgeschrittener unheilbarer Erkrankung. Notfall Rettungsmed. Mai 2021;24(3):203–10.

16. Adams HA, Salomon F. Entscheidungskonflikte am Notfallort. Anasthesiol Intensivmed Notfallmed Schmerzthe. Mai 2000;35(5):319–25.

17. Marung H, Wiese C. Palliativmedizin im Notarztdienst. Notf.med up2date. 10. September 2013;8(03):193–204.

18. Mühe K, Alt-Epping B, Duttge G, Steuer M, Zimmermann A. Die Minuten bis der “Doktor” kommt: Entscheidungsdilemmata von nichtärztlichem Rettungsfachpersonal. Palliativmedizin. Januar 2020;21(01):27–33.

19. Tong A, Sainsbury P, Craig J. Consolidated criteria for reporting qualitative research (COREQ): a 32-item checklist for interviews and focus groups. International Journal for Quality in Health Care. 16. September 2007;19(6):349–57.

20. Schade F, Rieder N, Jansky M, Nauck F. Online-Fokusgruppen mit haupt-und ehrenamtlich Tätigen im Setting der Hospiz-und Palliativversorgung. Zeitschrift für Palliativmedizin. Manuscript submitted for publication. 2023.

21. Pelz C, Schmitt A, Meis M. Knowledge Mapping as a Tool for Analyzing Focus Groups and Presenting Their Results in Market and Evaluation Research. Forum Qualitative Sozialforschung / Forum: Qualitative Social Research, [Internet]. 2004; Verfügbar unter: http://www.qualitative-research.net/index.php/fqs/article/view/601/1303.

22. Mayring Philipp. Qualitative Inhaltsanalyse Grundlagen und Techniken. Weinheim: Beltz; 2010.

23. Pulst A, Fassmer AM, Hoffmann F, Schmiemann G. Paramedics’ Perspectives on the Hospital Transfers of Nursing Home Residents—A Qualitative Focus Group Study. IJERPH. 26. Mai 2020;17(11):3778.

24. Bollig G, Brandt F, Ciurlionis M, Knopf B. Last Aid Course. An Education For All Citizens and an Ingredient of Compassionate Communities. Healthcare. März 2019;7(1):19.

25. Pinter G, Likar R, Schippinger W, Janig H, Kada O, Cernic K Herausgeber. Geriatrische Notfallversorgung [Internet]. Vienna: Springer Vienna; 2013 [zitiert 15. September 2022]. Verfügbar unter: http://link.springer.com/10.1007/978-3-7091-1581-7.

26. Redelsteiner C, Fohringer C, Ganaus P, Rottensteiner S, Hochsteger R, Weinert S, u. a. “RettungspflegerIn” – Erfahrungen einer interdisziplinären Berufsausbildung. In: Neumayr A, Baubin M, Schinnerl A Herausgeber. Zukunftswerkstatt Rettungsdienst [Internet]. Berlin, Heidelberg: Springer Berlin Heidelberg; 2018 [zitiert 15. September 2022]. S. 177–85. Verfügbar unter: http://link.springer.com/10.1007/978-3-662-56634-3_16.

27. Makowski C, Marung H, Callies A, Knacke P, Kerner T. Notarzteinsätze bei Palliativpatienten - Algorithmus zur Entscheidungsfindung und Behandlungsempfehlungen. Anästhesiol Intensivmed Notfallmed Schmerzther. 15. März 2013;48(02):90–6.

28. Nauck F. Ethische Aspekte in der Therapie am Lebensende. Med Klin. Oktober 2011;106(2):137–48.

29. Nauck F, Marckmann G, in der Schmitten J. Behandlung im Voraus planen – Bedeutung für die Intensiv-und Notfallmedizin. Anästhesiol Intensivmed Notfallmed Schmerzther. Januar 2018;53(01):62–70.

30. Stewart DW, Shamdasani P. Online Focus Groups. Journal of Advertising. 2. Januar 2017;46(1):48–60.

